# Mortality of Care Home Residents and Community-Dwelling Controls During the COVID-19 Pandemic in 2020: Matched Cohort Study

**DOI:** 10.1101/2021.04.24.21255968

**Authors:** Martin C Gulliford, A. Toby Prevost, Andrew Clegg, Emma Rezel-Potts

**Author notes:** Correspondence: Prof. Martin Gulliford, Address: SPHES, King’s College London, Guy’s Campus, London SE1 1UL.

## Abstract

**Objective:** To estimate mortality of care home (CH) residents, and matched community-dwelling controls, during the Covid-19 pandemic from primary care electronic health records.

**Design:** Matched cohort study

**Setting:** General practices contributing to the Clinical Practice Research Datalink Aurum Database in England.

**Participants:** There were 83,627 CH residents contributing data in 2020, with 26,923 deaths; 80,730 (97%) were matched on age, gender and general practice with 300,445 community-dwelling adults.

**Main outcome measures:** All-cause mortality. Adjusted rate ratios (RR) by negative binomial regression were adjusted for age, gender, number of long-term conditions (LTCs), frailty category, region, calendar month or week, and clustering by general practice.

**Results:** During April 2020, the mortality rate of CH residents was 27.2 deaths per 1,000 patients per week, compared with 2.31 per 1,000 for controls, RR 11.1 (95% confidence interval 10.1 to 12.2). Compared with CH residents, LTCs and frailty were differentially associated with greater mortality in community-dwelling controls. During April 2020, mortality rates per 1,000 patients per week for persons with 9+ LTCs were: CH, 37.9; controls 17.7; RR 2.14 (1.18 to 3.89). In severe frailty, mortality rates were: CH, 29.6; controls 5.1; RR 6.17 (5.74 to 6.62).

**Conclusions:** Individual-patient data from primary care electronic health records may be used to estimate mortality in care home residents. Mortality is substantially higher than for community-dwelling comparators and showed a disproportionate increase in the first wave of the Covid-19 pandemic. Multiple morbidity and frailty were associated with greater absolute risks but lower relative risks because community-dwelling frail or multi-morbid patients also experienced high mortality.

## INTRODUCTION

The Covid-19 pandemic had major impacts during 2020.^1^ The first wave of infections peaked during April 2020 in the UK, with more than 1,000 deaths per day within 28 days of a positive Covid-19 test. Transmission rates subsided during the summer months then rose again in the latter part of the year. In the second wave, with more widespread testing, the number of people in the UK with a positive COVID-19 test result peaked at 81,525 on 29^th^ December 2020.^1^ Early studies identified deprivation,^2^ household overcrowding,^3^ older age, male gender, obesity, comorbidity and ethnic minority status as being important risk factors for severe disease and mortality.^4^ Residents of care homes, which in the UK include residential homes providing support with personal care, and nursing homes providing support with personal care and assistance from qualified nurses, were severely affected by the pandemic. Contributing factors included the discharge of hospital patients to care homes with risks of disease transmission,^5^ limited availability of Covid-19 testing,^6^ limited supply of personal protective equipment^7^ and delayed development of guidance to ensure protection of the care home popualtion.^8^ Data from the Office for National Statistics showed that weekly counts of deaths of care home residents in England and Wales increased from 2,799 in the last week of February to 8,476 and 9,015 in the last two weeks of April 2020.^9^ Analysis of data reported to the Care Quality Commission (CQC) in England suggested that excess deaths represented about 6.5% of care home beds.^10^ Care home residents typically have multiple risk markers of vulnerability for severe Covid-19, but susceptibility to infection may also have been increased because the care home environment had potential to facilitate transmission of Covid-19 and outbreaks were frequent. However, rigorous epidemiological analysis has been limited. An editorial observed that ‘the covid-19 pandemic has placed a spotlight on how little is known about this sector, and the lack of easily accessible, aggregated data on the UK care home population’.^11^ In order to address this gap, we aimed to explore whether primary care electronic health records could be used to evaluate care home mortality during the pandemic. Jain et al.^12^ suggested that primary care electronic health records could be used to provide estimates for the living arrangement and care home residence of older adults that were comparable to those from census data. We aimed to use primary care electronic health records to estimate all-cause mortality of care home residents in comparison with matched community-dwelling controls in England during 2020. Since care home residence may be associated with greater vulnerability to adverse events, we evaluated frailty and multiple morbidity as possible effect modifiers.

## METHODS

### Data source and participant selection

The study drew on data from the CPRD Aurum database, a database of longitudinal primary care electronic health records in England.^13^ CPRD Aurum includes comprehensive records for symptoms, signs, medical diagnoses, tests and referrals, with data coded using Snomed CT.^13^ The CPRD Aurum database includes a total of 1,473 general practices in England with approximately 14.8 million registered patients at 1^st^ January 2020.

This study used data from the March 2021 release of CPRD Aurum. The initial data extract included 215,110 patients registered in CPRD Aurum general practices in England between 1^st^ January 2015 and 31^st^ December 2020 who were recorded as being resident in a care home. We employed a list of 49 medical codes indicative of care home residence. The most frequently recorded index care home codes were ‘lives in a nursing home [or] care home’ (Supplementary Table 1). There were 28,531 (13%) patients with index codes of ‘patient died in a nursing home [or] care home’. For these, patients we assumed that they were resident in the care home for 90 days before death. The median length of stay is two years for care home residents, and one year for nursing home residents,^14^ but we assumed that patients with first codes for ‘died in nursing/care home’ would have lower than average lengths of stay. In sensitivity analyses, we found that varying this assumed duration between 14 and 365 days had negligible influence on estimates. For each patient, the start date was the latest of the patient’s start of registration or the first care home code. The end of the patient’s record was the earliest of the end of patient registration, the death date recorded by CPRD and the last data collection date for the practice. We included patients aged 18 to 104 years of age.

For 83,627 care home residents contributing person-time during 2020, a matched comparison cohort of community-dwelling adults was sampled from the list of all patients registered in the CPRD Aurum March 2021 release after excluding care home residents. Control patients were matched for general practice, gender and year of birth, and had a start date that was no later than 18 months after the start date for matched cases. Up to four community-dwelling control participants were randomly sampled with replacement^15^ for each care home resident. Care home residents were omitted from this analysis where there were no eligible matched controls. As the difference in mortality between care home residents and community-dwelling controls was found to be greater than anticipated, control selection, data extraction and data analysis were repeated to confirm the reproducibility of findings.

### Main measures

The primary measure of interest was mortality from any cause based on the CPRD death date. Death records were included up to seven days after the end of record to allow for possible delayed recording into primary care records. Covariates were age, gender, region in England, multiple morbidity and frailty category. Age in 2020 and was divided into the age-groups of 18 to 64, 65 to 74, 75 to 84, 85 to 94 and 95 to 104 years. Multiple morbidity was represented by a count of 20 conditions, ever recorded in each patient’s record up to the end of 2020, from the list of atrial fibrillation, cancer, chronic kidney disease, chronic obstructive pulmonary disease, dementia, depression, diabetes mellitus, epilepsy, frailty fractures, heart failure, haemorrhagic stroke, hypertension, ischaemic heart disease, ischaemic stroke, other mental health diagnoses, peripheral arterial disease, palliative care, rheumatoid arthritis or transient ischaemic attack. Frailty was evaluated from coded records of deficits noted in CPRD Observation files according to the e-Frailty index, as described by Clegg et al.^16^ Coded records of frailty index scores and frailty index categories were also analysed to inform frailty classification with the highest recorded value being employed. The categories of ‘non-frail’, ‘mild frailty’, ‘moderate frailty’ and ‘severe frailty’ were employed for analysis.

### Statistical analysis

We initially analysed eligible care home patient records between 1^st^ January 2015 and 31^st^ December 2020. We divided records into calendar months, calculating the number of deaths and person time at risk for each month. We fitted a negative binomial regression model using data up to the end of 2019 as the training dataset, with counts of observed deaths as dependent variable and age-group, gender, region, multiple morbidity, frailty category, calendar month and calendar year as predictors. Month was fitted as a factor, while year was fitted as a continuous predictor. Multiple morbidity was fitted as a factor with categories from one to nine or more morbidities, with a separate category for ‘none recorded’. Frailty category was fitted as a factor. Robust standard errors were employed to allow for general practice clustering, but the general practice effect was allowed to differ between care home residents and controls because the former might also have been clustered within care homes. From the fitted model, we estimated predicted deaths by month for the period 2015 to 2020. We compared predicted and observed deaths graphically. In order to evaluate mortality in 2020 in more detail, we analysed data for care home residents and community controls, evaluating counts of deaths and persons at risk by calendar week. We fitted a negative binomial model, with robust standard errors, now including interaction terms that allowed the associations of LTCs and frailty with mortality to differ between care home residents and community controls. Observed and predicted mortality rates were plotted for each level of multi-morbidity and frailty. Finally, to summarise the results we fitted models separately for the periods of January to March, April, and May to December. Analyses were performed using the ‘statsmodels’^17^ package in Python 3.8.3. The ‘matplotlib’^18^ package in Python and the ‘ggplot2’^19^ package in the R program were used for data visualization.

## RESULTS

There were 215,110 patients who were registered at general practices in England and were recorded as resident in a Care Home, who contributed follow-up between 1^st^ January 2015 and 31^st^ December 2020. There were 137,024 (64%) women; 97,192 (45%) were aged 85 to 94 years and 24,685 (11%) were aged 95 or older; 180,390 (84%) had two or more morbidities. Figure 1 shows the distribution of observed deaths (red line) by month from 2015 to 2020, compared with predicted values estimated from 2015 to 2019 data (blue line). It was clear that there was a substantial excess of observed over predicted deaths in early 2020, with a peak in April 2020.

**Figure 1:**
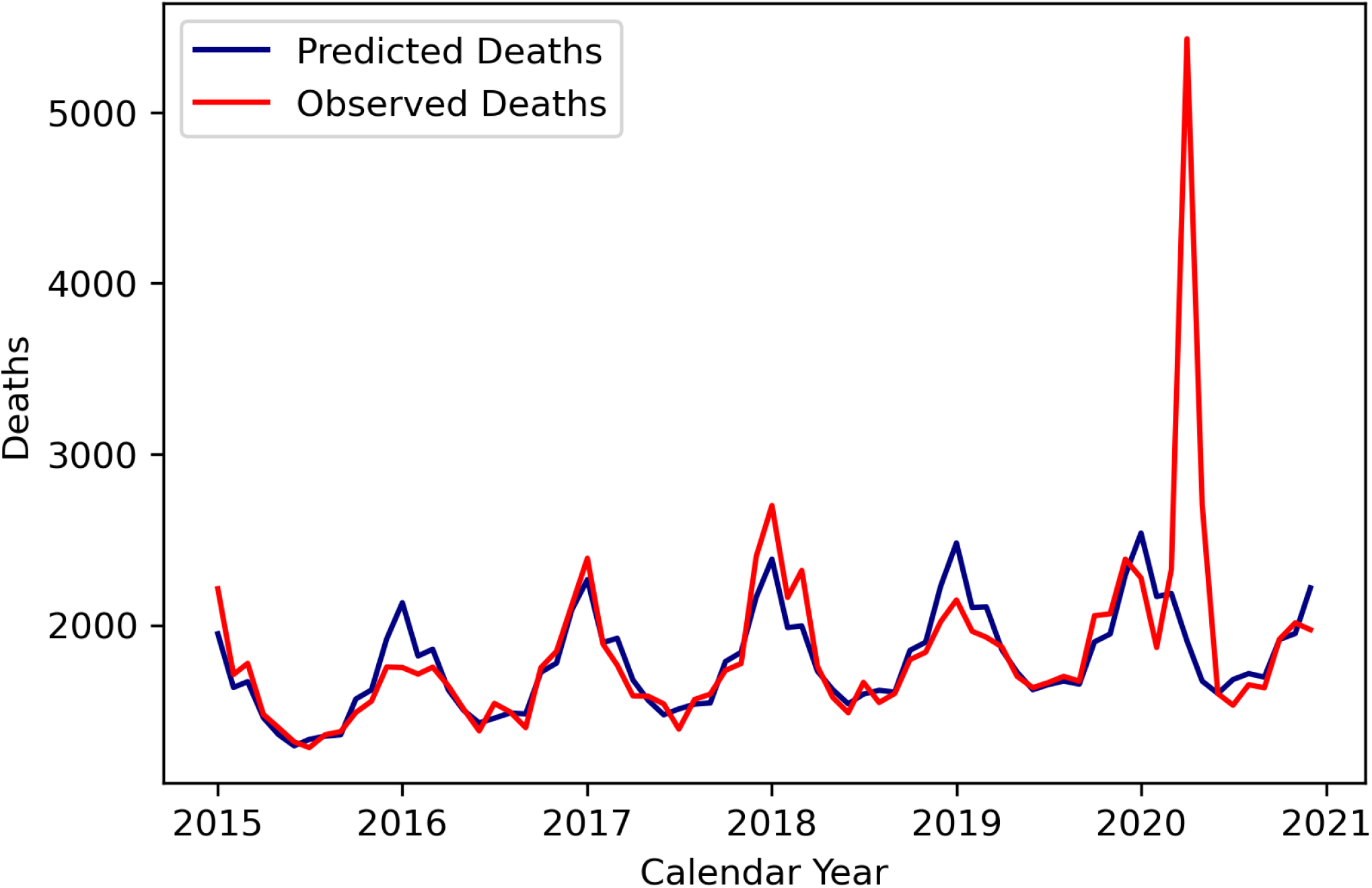
Monthly counts of observed deaths of care home residents between 2015 and 2020 (red) with predicted deaths from Poisson model fitted to 2015 to 2019 data (blue).

In order to investigate pandemic peak in mortality in more detail, analyses were then restricted to 83,627 care home residents who were registered during 2020, of whom 80,730 (97%) were matched with 300,445 community-dwelling controls. Characteristics of the sample are shown in Table 1. Care home residents and community controls were similar with respect to matching variables of gender and age-group, but care home residents generally showed higher counts of long-term conditions and greater levels of frailty.

**Table 1:** Characteristics of participants. Figures are frequencies (percent of column total).

| Variable | Category | Care Home Residents | Community Controls |
| --- | --- | --- | --- |
| **Total** |  | 80,730 | 300,445 |
| **Gender** | Female | 51,873 (64) | 191,523 (64) |
|  | Male | 28,857 (36) | 108,922 (36) |
| **Age-group (years)** | 18 to 64 | 9,250 (11) | 36,801 (12) |
|  | 64 to 74 | 8,234 (10) | 32,699 (11) |
|  | 74 to 84 | 20,777 (26) | 81,402 (27) |
|  | 84 to 94 | 34,277 (42) | 126,977 (42) |
|  | 94 to 104 | 8,190 (10) | 22,563 (8) |
| **Number of LTCs** | 1 | 10,466 (13) | 64,481 (21) |
|  | 2 | 14,264 (18) | 64,935 (22) |
|  | 3 | 16,199 (20) | 51,240 (17) |
|  | 4 | 14,448 (18) | 33,622 (11) |
|  | 5 | 10,084 (12) | 18,773 (6) |
|  | 6 | 5,986 (7) | 8,996 (3) |
|  | 7 | 3,052 (4) | 3,849 (1) |
|  | 8 | 1,288 (2) | 1,387 (0) |
|  | 9+ | 614 (1) | 560 (0) |
|  | None recorded | 4,349 (5) | 52,602 (18) |
| **Frailty Category** | Non-frail | 5,284 (7) | 61,829 (21) |
|  | Mild | 13,467 (17) | 82,645 (28) |
|  | Moderate | 21,400 (27) | 75,600 (25) |
|  | Severe | 40,367 (50) | 69,337 (23) |
|  | Not recorded | 212 (0) | 11,034 (4) |

Analyses were repeated using calendar weeks for analysis, with data presented as deaths per 1,000 patients per week by age-group and gender (Figure 2, upper panel). There was a peak in observed deaths between 6th April 2020 and 26th April 2020. Mortality rates were higher in men than women and increased in successive age-groups. The highest age-specific mortality rate was 63.6 (95% confidence interval 63.1 to 64.1) per 1,000 patients per week in men aged 95-104 years between 13th-19th April. Excess deaths, calculated as the difference between observed and predicted deaths, were summed across all age-groups (Figure 2, lower panel). Across all ages, excess deaths peaked in men between 13th-19th April, at 27.7 per 1,000 per week.

**Figure 2:**
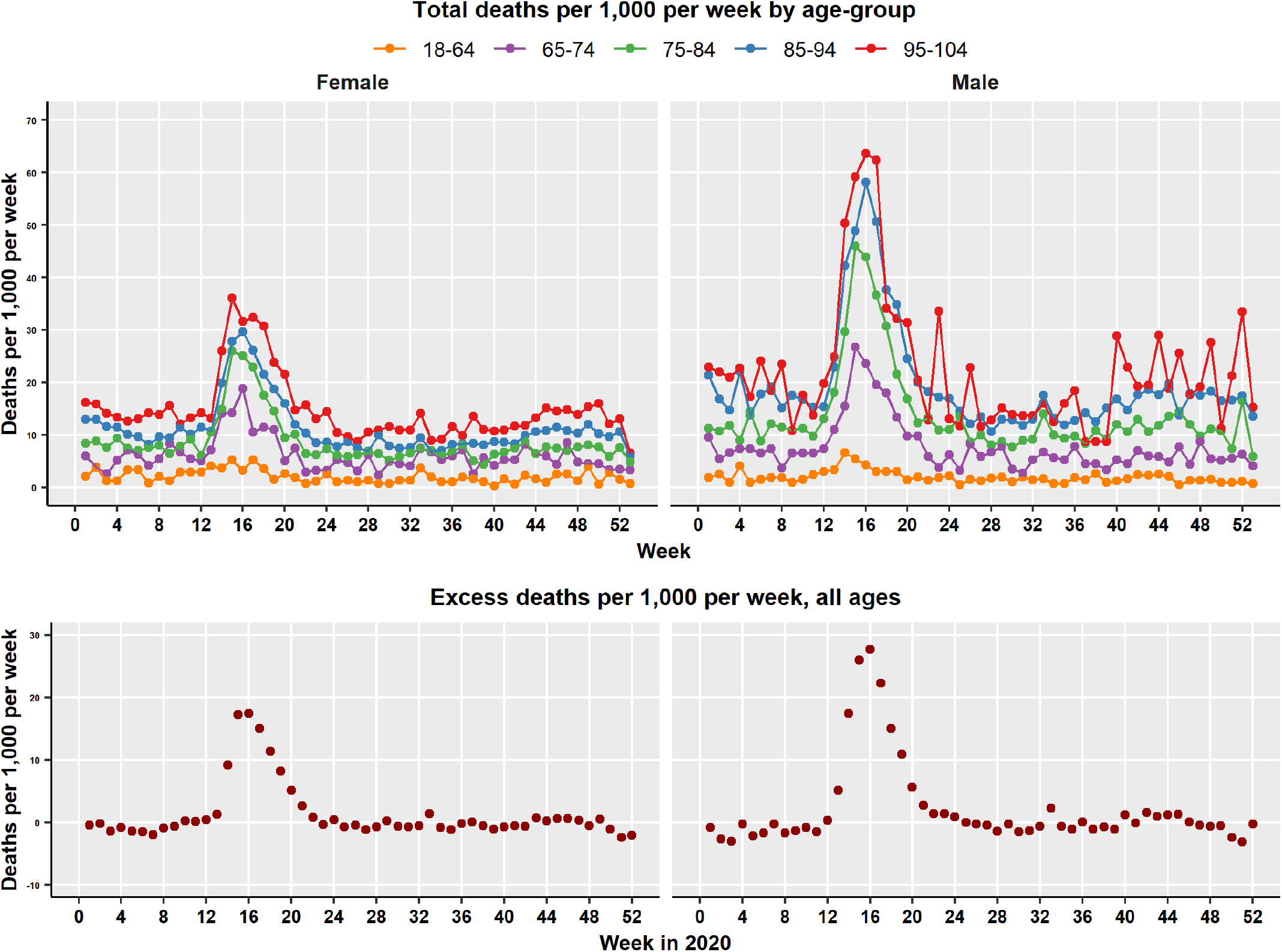
Total deaths per 100 patients per week during 2020 by age-group and gender (upper panel). Excess deaths (observed minus predicted) across all ages per 100 patients per week (lower panel).

Figure 3 shows mortality rates per 1,000 patients per week for each week of 2020 for care home residents (red) and community controls (blue). Data are presented separately by number of long-term conditions (upper panel) and frailty category (lower panel). Points represent crude mortality rates, while solid lines represent mortality predicted from the adjusted Poisson model fitted to 2020 data. There was a peak in observed deaths between 6^th^ April 2020 and 26^th^ April 2020 that was evident in both care home residents and community controls. Mortality of care home residents was always higher than for community controls. Mortality also increased with number of long-term conditions and frailty category. However, the effect of increasing long-term condition count or frailty category was greater for community controls than for care home residents. Community-dwelling patients with higher long-term condition counts or severe frailty showed mortality that was more similar to care home residents than those with fewer long-term conditions or who were non-frail.

**Figure 3:**
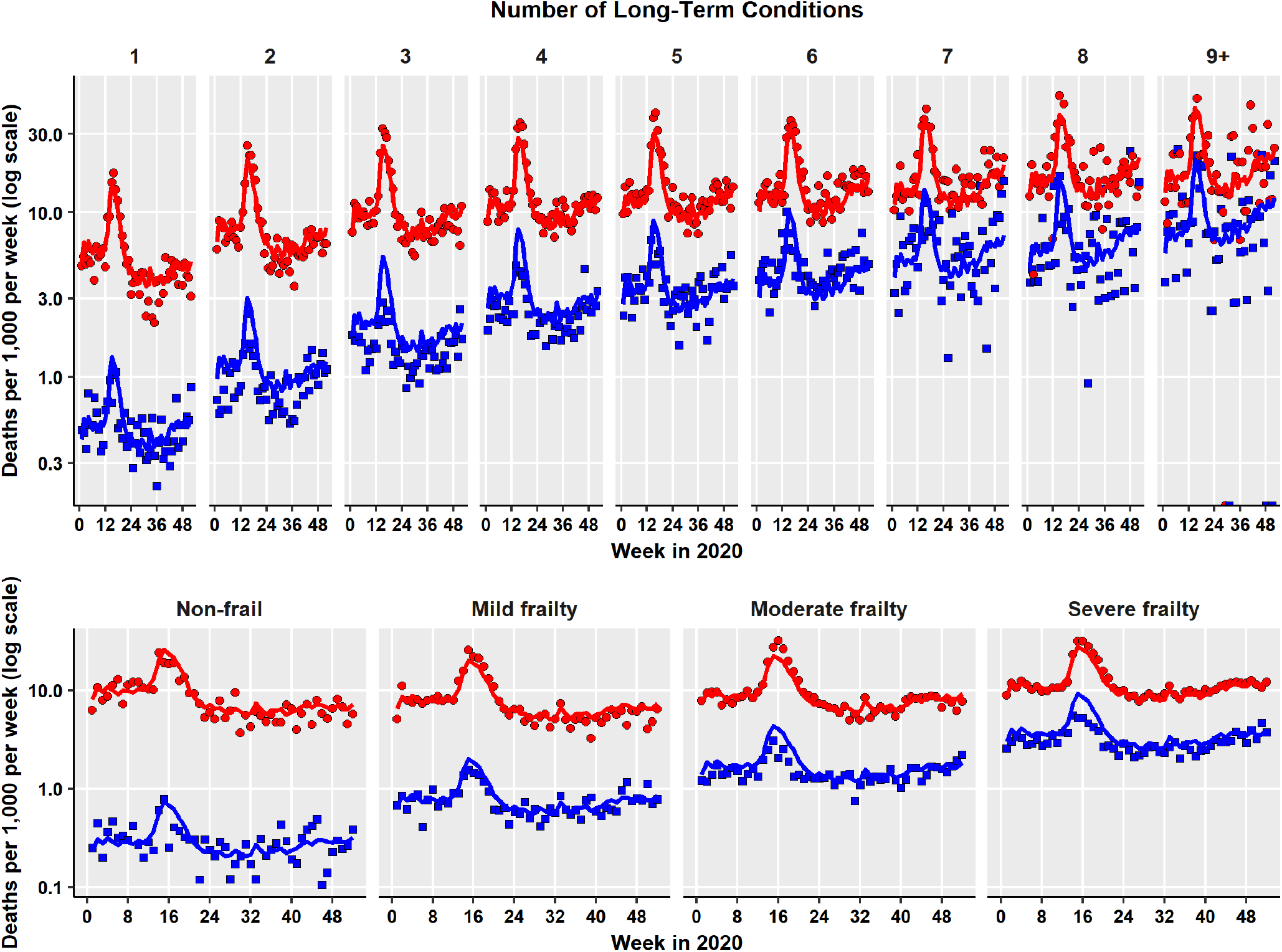
Total deaths per 100 patients per week during 2020 by number of long-term conditions (upper panel) and frailty category (lower panel). Red symbols, care homes residents; blue symbols, community dwelling controls. Points represent crude rates, lines represent predictions from adjusted regression model.

Table 2 shows data aggregated for the periods January to March, April, and May to December 2020. Mortality of care home residents was higher in the April period than the other periods; this increase was evident at each level of frailty with absolute risks of mortality increasing with frailty level. Mortality of community controls was also higher in April as compared to the other periods; the increase was proportionately smaller than for care home residents but, in absolute terms, the increase was greatest for patients with the most advanced level of frailty. Comparing care home residents and community controls, the adjusted relative mortality rate decreased with increasing level of frailty, reflecting the higher mortality of frail community controls. However, relative risks were higher in April period than in other periods of 2020.

**Table 2:**
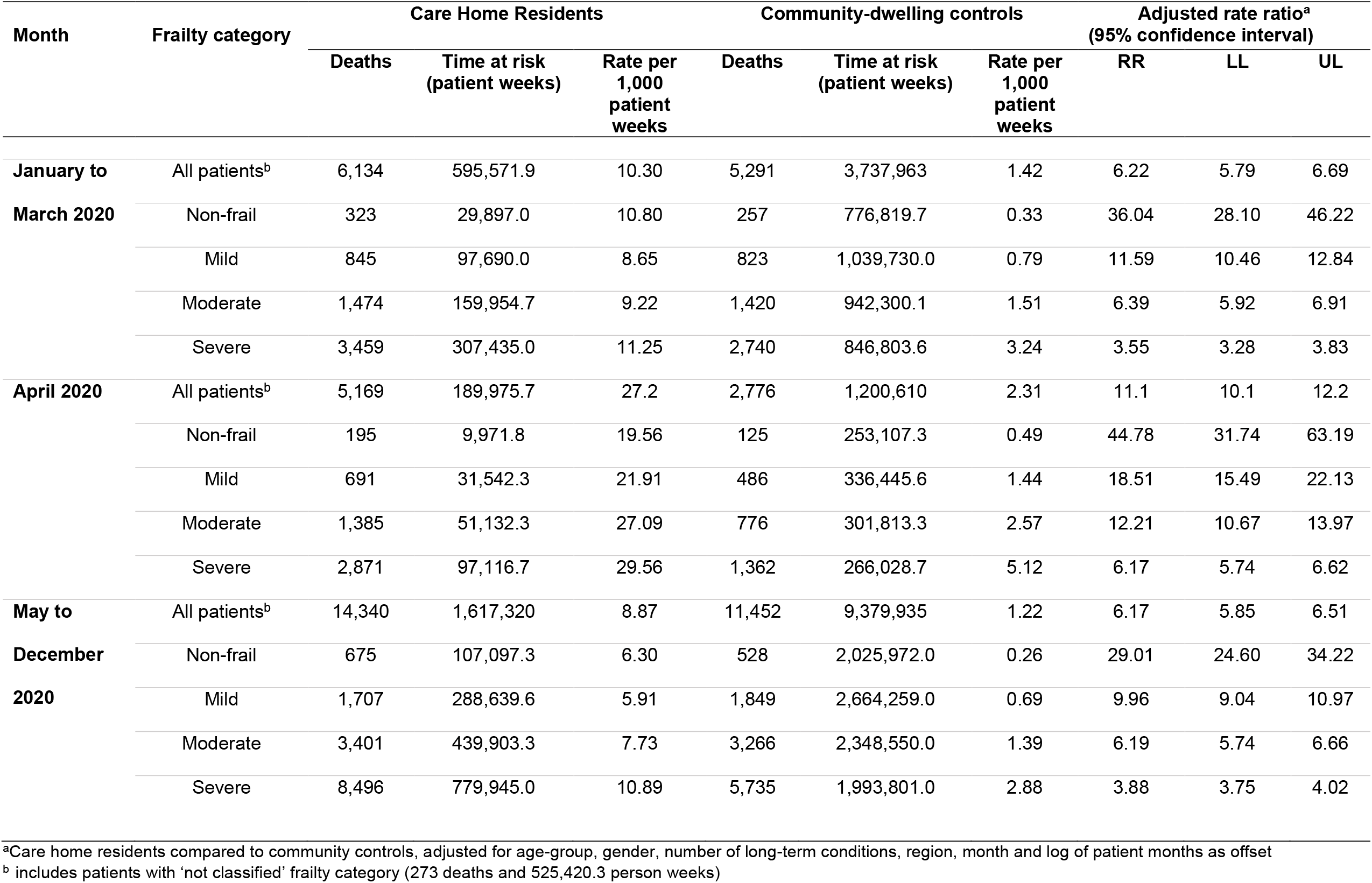
Deaths and person-time for care home residents and matched controls by year. RR, adjusted rate ratio; LL and UL, lower and upper bounds of 95% confidence interval.

| Month | Frailty category | Care Home Residents |  |  | Community-dwelling controls |  |  | Adjusted rate ratio<sup>a</sup><br>(95% confidence interval) |  |  |
| --- | --- | --- | --- | --- | --- | --- | --- | --- | --- | --- |
|  |  | Deaths | Time at risk<br>(patient weeks) | Rate per<br>1,000<br>patient<br>weeks | Deaths | Time at risk<br>(patient weeks) | Rate per<br>1,000<br>patient<br>weeks | RR | LL | UL |
| **January to** | All patients<sup>b</sup> | 6,134 | 595,571.9 | 10.30 | 5,291 | 3,737,963 | 1.42 | 6.22 | 5.79 | 6.69 |
| **March 2020** | Non-frail | 323 | 29,897.0 | 10.80 | 257 | 776,819.7 | 0.33 | 36.04 | 28.10 | 46.22 |
|  | Mild | 845 | 97,690.0 | 8.65 | 823 | 1,039,730.0 | 0.79 | 11.59 | 10.46 | 12.84 |
|  | Moderate | 1,474 | 159,954.7 | 9.22 | 1,420 | 942,300.1 | 1.51 | 6.39 | 5.92 | 6.91 |
|  | Severe | 3,459 | 307,435.0 | 11.25 | 2,740 | 846,803.6 | 3.24 | 3.55 | 3.28 | 3.83 |
| **April 2020** | All patients<sup>b</sup> | 5,169 | 189,975.7 | 27.2 | 2,776 | 1,200,610 | 2.31 | 11.1 | 10.1 | 12.2 |
|  | Non-frail | 195 | 9,971.8 | 19.56 | 125 | 253,107.3 | 0.49 | 44.78 | 31.74 | 63.19 |
|  | Mild | 691 | 31,542.3 | 21.91 | 486 | 336,445.6 | 1.44 | 18.51 | 15.49 | 22.13 |
|  | Moderate | 1,385 | 51,132.3 | 27.09 | 776 | 301,813.3 | 2.57 | 12.21 | 10.67 | 13.97 |
|  | Severe | 2,871 | 97,116.7 | 29.56 | 1,362 | 266,028.7 | 5.12 | 6.17 | 5.74 | 6.62 |
| **May to** | All patients<sup>b</sup> | 14,340 | 1,617,320 | 8.87 | 11,452 | 9,379,935 | 1.22 | 6.17 | 5.85 | 6.51 |
| **December 2020** | Non-frail | 675 | 107,097.3 | 6.30 | 528 | 2,025,972.0 | 0.26 | 29.01 | 24.60 | 34.22 |
|  | Mild | 1,707 | 288,639.6 | 5.91 | 1,849 | 2,664,259.0 | 0.69 | 9.96 | 9.04 | 10.97 |
|  | Moderate | 3,401 | 439,903.3 | 7.73 | 3,266 | 2,348,550.0 | 1.39 | 6.19 | 5.74 | 6.66 |
|  | Severe | 8,496 | 779,945.0 | 10.89 | 5,735 | 1,993,801.0 | 2.88 | 3.88 | 3.75 | 4.02 |

<sup>a</sup>Care home residents compared to community controls, adjusted for age-group, gender, number of long-term conditions, region, month and log of patient months as offset
<sup>b</sup> includes patients with 'not classified' frailty category (273 deaths and 525,420.3 person weeks)

Figure 4 presents a forest plot of the adjusted mortality rate ratios, comparing care home residents with community controls, by period and number of long-term conditions and frailty level. Compared with community controls, the relative risk of mortality for care home residents decreased with advancing number of long-term conditions or advancing frailty category. At each level of morbidity or frailty, the relative risk of mortality for care home residents was higher during April compared with January to March or May to December.

**Figure 4:**
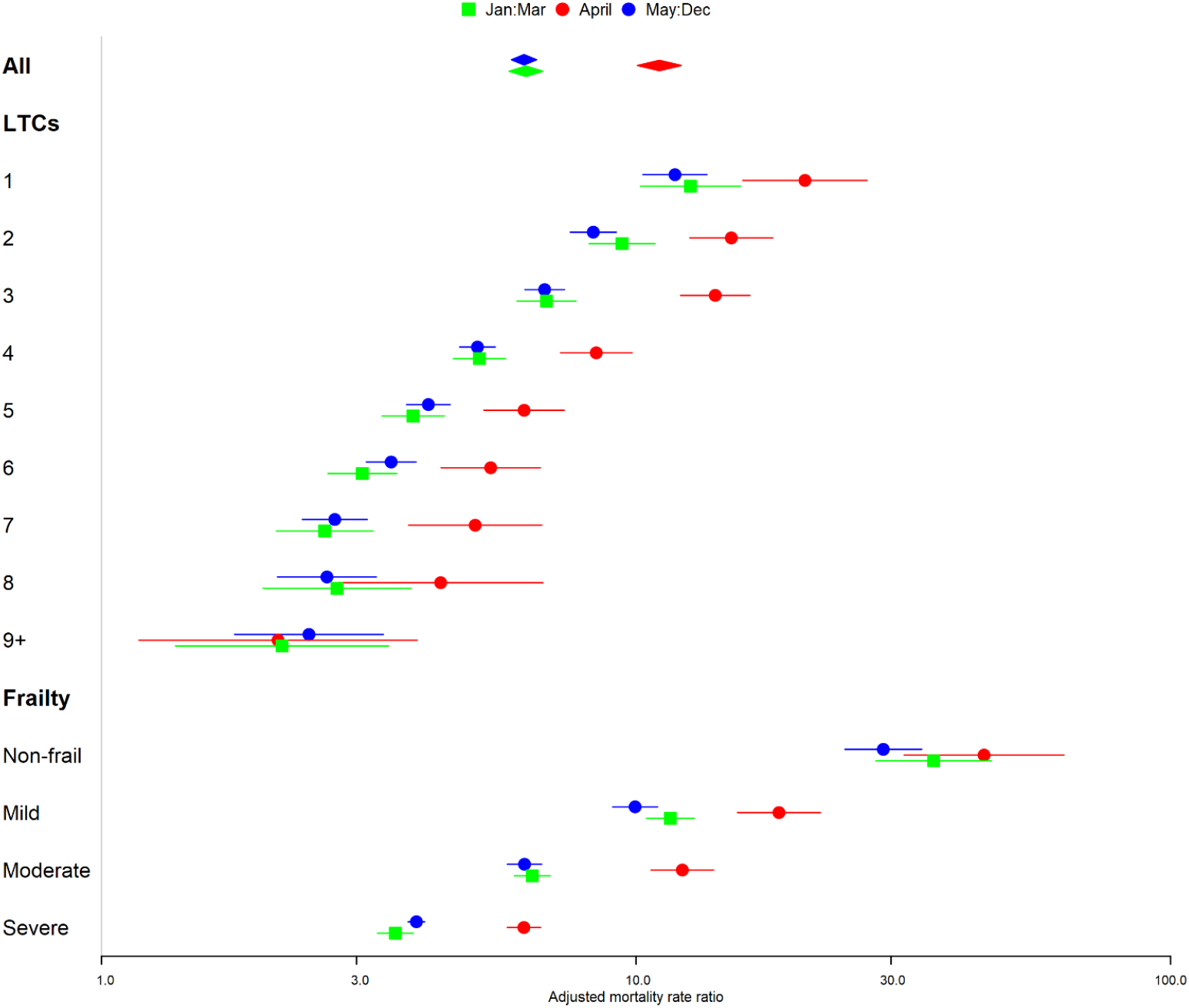
Forest plot showing adjusted mortality rate ratios, comparing care home residents and community controls, at each level number of long-term conditions and frailty category. January to March 2020, green; April 2020, red; May to December 2020, green.

## DISCUSSION

### Main findings

The first wave of the Covid-19 pandemic is acknowledged to have had a particularly severe impact on patients living in care homes. However, there has been a lack of patient-level data concerning the health outcomes of care home residents during the pandemic.^11^ This analysis shows that primary care electronic health records have potential to provide timely and relevant information concerning the care home population. Analyses quantified the first wave of Covid-19 mortality in April 2020 and showed that mortality peaked between 6^th^ and 26^th^ April, being strongly associated with advanced age, male gender, multiple morbidity and frailty category. Compared with community-dwelling control patients, mortality for care home residents was four to five times higher before the onset of the pandemic. Care home residents were disproportionately affected and during the month of April 2020 after allowing for differences in case-mix, mortality of care home residents was more than ten times higher than for community-dwelling patients overall. The level of frailty and number of long-term conditions were found to be effect modifiers, being more strongly associated with mortality of community-dwelling patients than those living in care homes. Frail patients living in the community might have been exposed to similar risks of Covid-19 transmission to care home patients if they were supported by carers or care providers who might have had possibly limited or inadequate protections. Conversely, residence in a care home was associated with greater mortality even in the minority of patients who were classified as non-frail. This might to some extent be explained by data recording limitations, as care home admission was often associated with discontinuities of general practice registration that could be associated with incomplete medical records. However, the shared environment of the care home might be associated with heightened mortality risk irrespective of frailty category.

### Strengths and Limitations

An important strength of this study was the use of longitudinal health records to estimate predicted mortality during 2020 based on data for the preceding five years, taking into account differences in age and gender distribution, morbidities, region, calendar month and secular trends over years. This enabled us to quantify excess deaths during the pandemic months. We were also able to draw on a matched population-based comparison cohort to quantify changes in the relative risk of mortality in care homes during the pandemic after adjusting for covariates. We drew on a well-described database,^13^ and the quality of data offered by electronic health records has been shown to be generally high.^20^ However, we acknowledge that there could be misclassification of care home status and it is possible that care home residence might be under-recorded. This might lead to the care home sample lacking representativeness if, for example, care home residence is only recorded for those residents with greater need for general practice services. Selection bias and misclassification of care home status might have the effect of either reducing or inflating estimated associations. We included a count of important long-term conditions as well as analysing frailty category. In the cumulative deficit model, frailty and multiple morbidity are closely related concepts^21^ but more accurate phenotypic characterisation of patients frailty status over time would have added to the study.^22^ Deprivation is associated with reduced healthy life expectancy, which could lead to care home admission. Patients were matched for general practice, so it was not possible to adjust for deprivation at the general practice-level. We did not employ individual postcode-level deprivation scores as these might have presented difficulties if the care home postcode did not reflect deprivation exposures over the life-course. Control sampling was with replacement and duplicated controls were included to reduce bias.^15^

### Comparison with other studies

Previous studies of care home mortality during the Covid-19 pandemic have mainly drawn on data from care home records.^10 23^ Morciano et al.^10^ analysed data for numbers of deaths reported to the care quality commission and estimated that over the first seven months of 2020, deaths accounted for 6.5% of care home beds. The estimates from our analyses are not directly comparable because we estimated the mortality rate per 1,000 residents per week. Dutey-Magni et al.^23^ analysed data collected by care homes for incidence of Covid-19 and mortality. Their findings, like our study, suggested that deaths were frequent among residents who were probably infected with SARS-CoV-2 but were not tested. Burton et al.^24^ found that outbreaks of Covid-19 were frequent within care homes and most deaths occur in the context of outbreaks.^10 24^ We did not have data to identify individuals at the same care homes and the possible clustering of deaths at care homes could not be investigated in our data. Hollinghurst et al.^25^ analysed linked primary care and administrative records for the population of Wales and found that care homes showed increased mortality during the first wave of the pandemic. Their estimates were generally lower than we present here. However, their analysis using the Cox model could be associated with non-proportional hazards because analysis time encompassed a period when risks were changing daily. We estimated adjusted relative risks for each week of the pandemic and showed that there was a substantial increase in the relative risk of mortality associated with care home residence during the first wave. Other studies confirm that background mortality is very high in care home residents. Vossius et al.^26^ found that annual mortality of nursing home residents was 31.8%. Shah et al.^27^ analysing the THIN primary care database for 2009 found that the age and sex standardised mortality ratio for nursing home residents was 419 and for residential home residents was 284, consistent with the elevated relative rates observed in the present analyses.

### Implications

Despite awareness since the very early stages of the UK pandemic of evidence from other contexts that Covid-19 severity was likely to be greatest among the elderly,^28^ there were delays in policy guidance which correspond with the timing of Covid-19 mortality peaks in care home observed in April 2020. In the event of future transmission increases, earlier implementation of testing and isolation strategies and greater consideration of the effects of hospital discharge to care homes may be crucial. This study has also highlighted that there is heterogeneity in the care home population, indicating that the most elderly males may require particular protection or shielding during periods of high transmission. Regional variations in mortality might also indicate that more localised approaches should be explored. Further assessment is required of longer-term issues that have may have contributed to higher rates of care home Covid-19 mortality such as decreases in local authority social care spending since 2010, increased privatisation,^29^ staff shortages ^30 31^ and the lack of integration of health and social care services.^32^ The high mortality of care home residents during non-pandemic months, even after allowing for the level of morbidity, might be accounted for by admissions for end-of-life care. Nevertheless, prospective investigations of mortality in care homes are justified.

## Conclusions

Individual-patient data from primary care electronic health records may be used to estimate mortality in care home residents either in comparison with non-pandemic periods or with population controls. Analyses confirmed the disproportionate impact of the first-wave of the Covid-19 pandemic on the care home population, especially in comparison with community-living comparators. Estimation of deaths per calendar week were mapped against delays in action to isolate care home residents. In the event of further outbreaks, this study provides evidence for earlier, decisive action to protect these vulnerable populations.

## Data Availability

Data sharing requests should be sent to. Data release is subject to approval from CPRD.

## Ethical Approval

The protocol for the study was approved by the CPRD Independent Scientific Advisory Committee protocol number 20_000214.

## Data sources

The study is based in part on data from the Clinical Practice Research Datalink obtained under license from the UK Medicines and Healthcare products Regulatory Agency. However, the interpretation and conclusions contained in this report are those of the authors alone. Requests for data sharing should be directed to the corresponding author.

## Contributorship

MG wrote the study protocol and conducted data analyses; ERP provided advice on study design, data analysis and interpretation. ATP advised on statistical modelling and interpretation of results. AC advised on frailty classification and interpretation of results. All authors contributed to drafting the paper and approved the final draft. MG is guarantor.

## Transparency

The lead author (MG) affirms that the manuscript is an honest, accurate, and transparent account of the study being reported; that no important aspects of the study have been omitted; and that any discrepancies from the study as originally planned (and, if relevant, registered) have been explained.

## Data sharing

Data sharing requests should be sent to. Data release is subject to approval from CPRD.

---

**Supplementary Table 1.** First recorded medical codes for care home residence for 215,110 patients registered from 2015 to 2020 (14 most frequent codes shown).

| Medical Term | Frequency |
| --- | --- |
| Lives in a nursing home | 78,897 |
| Lives in care home | 54,217 |
| Patient died in nursing home | 25,652 |
| Care home visit | 9,383 |
| Seen in nursing home | 8,198 |
| Care home visit for initial patient assessment | 6,724 |
| Nursing home visit note | 5,715 |
| Care home enhanced services administration | 5,054 |
| Care home visit for follow-up patient review | 4,097 |
| Weekly care home ward round | 3,509 |
| Patient died in care home | 2,879 |
| Admission to nursing home | 2,717 |
| Nursing home | 2,642 |
| Discharge to nursing home | 1,312 |
| All other codes | 4,114 |

